# Twins in Guinea-Bissau have a ‘thin-fat’ body composition compared to singletons

**DOI:** 10.1101/2021.07.07.21260161

**Authors:** Rucha Wagh, Morten Bjerregaard-Andersen, Souvik Bandyopadhyay, Pranav Yajnik, Rashmi B Prasad, Suhas Otiv, Stine Byberg, Ditte Egegaard Hennild, Gabriel Marciano Gomes, Kaare Christensen, Morten Sodemann, Dorte Møller Jensen, Chittaranjan Yajnik

## Abstract

‘Thrifty phenotype’ hypothesis proposed that fetal undernutrition increases risk of diabetes in later life. Undernourished low birthweight Indian babies are paradoxically more adipose compared to well-nourished European babies, and are at higher risk of diabetes in later life. Twin pregnancies are an example of *in utero* growth restrictive environment due to shared maternal nutrition. There are few studies of body composition in twins. We performed secondary analysis of anthropometric body composition of twins and singletons in Guinea-Bissau, an economically deprived African country.

Anthropometric data was available on 7–34 year-old twins (n=209, 97 males) and singletons (n=182, 86 males) in the Guinea-Bissau Twin Registry at the Bandim Health Project. Twins had lower birth weight (2420 vs 3100 g, p<0.001); and at follow-up, lower height (HAZ mean Z-score difference, -0.21, p=0.055), weight (WAZ -0.73, p=0.024) and BMI (BAZ -0.22, p=0.079) compared to singletons but higher adiposity (skinfolds: +0.33 SD, p=0.001). Twins also had higher fasting (+0.38 SD, p<0.001) and 2-hr OGTT glucose concentrations (+0.29 SD, p<0.05). Linear mixed-effect model accounting for intrapair correlations and interactions confirmed that twins were thinner but fatter across the age range. Data on maternal morbidity and prematurity were not available in this cohort.

African populations are known to have a muscular (less adipose) body composition. Demonstration of a thin-fat phenotype in twins in a low socioeconomic African country supports the thesis that it could be a manifestation of early life undernutrition and not exclusive to Indians. This phenotype could increase risk of diabetes and related conditions.

## INTRODUCTION

The ‘thrifty phenotype’ hypothesis proposes that adult type 2 diabetes and related metabolic traits are a result of the fetus having to be thrifty in managing its nutrition during intrauterine life^1^. The initial report in an English study showed that prevalence of type 2 diabetes was higher in those with lower birth weight^2^. This association has been replicated in other populations ^3^. While the exposures in the original thrifty phenotype referred to low birth weight alone, this has since been extended to include relative abnormalities in body proportions, body composition, and development of organs and physiological-endocrine systems ^4^. India has a high prevalence of type 2 diabetes as well as intra-uterine undernutrition; this situation helped expand the thrifty phenotype concept. The Pune Maternal Nutrition Study showed that the short and thin (low ponderal index) Indian newborn weighing only 2.7 kg had more subcutaneous and visceral fat compared to a 3.5 kg English baby, along with higher concentrations of insulin and leptin and lower concentrations of adiponectin in cord blood ^5,6,7^. Since these differences were already seen at birth, the ‘thin-fat’ phenotype could be at least partially ascribed to the influence of intrauterine undernutrition on body composition of the developing fetus (nutritional programming). This is possibly achieved through epigenetic mechanisms regulating fetal growth and development ^8^. ‘Thin-fat’ phenotype persists through childhood, adolescence and adult age, and is associated with an increased risk of diabetes ^9, 10,11,12^. Such a phenotype (albeit milder) is also described in South-East Asian populations ^13^ but rarely investigated in other populations.

Twins are born with lower birth weight as compared to singletons and therefore lend themselves to DOHaD research. Comparison of adult disease risk in birthweight-discordant twins, especially monozygotic (MZ), is valuable as it allows the dissection of genetic and intra-uterine epigenetic influences. The smaller of the twins (irrespective of zygosity) have a higher risk of developing type 2 diabetes ^14^. Comparison between twins and singletons also provides an opportunity to study the effects of relative intra-uterine undernutrition. Most of the studies have concentrated on lower birthweight alone, and adiposity has been under-explored in twins as compared to singletons. These studies have been done in relatively well-nourished Western populations. It has been discussed that the biology of small size is different in twins from that in singletons and the difficulties in interpretation of association of birth size with later cardiometabolic risk in twins have been highlighted ^15^. Studying twins in low socioeconomic populations could be more informative, as twinning would compound the effects of maternal undernutrition in pregnancy with the ‘distributive’ biology of twin growth (30 percent of monozygotic and all dizygotic twins are dichorionic^16^). Thus, twin studies have the potential to reveal outcomes which could otherwise be missed. In Africa, there is a high natural twining rate, including in Guinea-Bissau, a low-income country ^17,18^. A twin registry was set up by the Bandim Health Project (BHP, www.bandim.org) in the capital city of Bissau^19^. Follow-up of twins in this registry showed that young twins had higher plasma glucose concentrations both in fasting and in the post-glucose state compared to age-matched singletons^20,21^. This study measured skinfolds in addition to the routine anthropometry and provided an opportunity to compare adiposity of twins with that of singletons. We hypothesized that twins in Guinea-Bissau would be thinner but fatter compared to singletons.

## METHODS

### Cohort description

This is a secondary analysis of previously collected (2009-2012) data from one of Africa’s first twin registries set up by the Bandim Health Project (BHP) in Guinea-Bissau ^19^. The BHP is a health and demographic surveillance site (HDSS), conducting epidemiological and health related research in the capital Bissau over the past 40 years. The results of the two main metabolic twin studies have previously been reported ^20,21^. Briefly, twins (n=209) and singleton controls (n=182), identified randomly from the HDSS register between 7-34 years of age were investigated for risk of diabetes and metabolic syndrome with a weight-adjusted oral glucose tolerance test (OGTT) (1.75 g glucose/kg body weight, maximum 75 g). The majority of measurements were done by a HemoCue Glucose 201+ apparatus (HemoCue, Ängelholm, Sweden), which uses capillary blood and then automatically converts the results to plasma values. In 7 % of the twins and 21 % of the singletons an AccuChek Active apparatus (Roche Diagnostics, Indiana, USA) was used, due to shortage of HemoCue cuvettes. Other biochemical measurements included total and HDL cholesterol, and triglycerides by standard enzymatic methods. Anthropometric measurements were performed by trained personnel. They included: height, weight, mid-upper arm circumference (MUAC), waist and hip circumferences using standard techniques; Skinfolds (Triceps, biceps, subscapular and suprailiac) were measured using a Harpenden Skinfold Caliper (Baty International, West Sussex, United Kingdom).

Deliveries happened at home or in local institutions. Birthweight was available only in a limited number (n=102) who were delivered in a hospital or health institution. Other anthropometric measurements (skinfolds, length and circumferences) were not available at birth.

### Statistical analysis

The primary analysis was to compare anthropometric measurements of height, thinness-obesity (BMI) and adiposity (skinfolds) between twins and singletons (Table 1). At the time of measurements, the age range was quite wide (7-34 years), therefore our preliminary analysis compared body size and composition based on sex specific WHO Z scores which provide data from birth to 19-years. The Nineteen-year value was used as reference to calculate Z -scores for older subjects.

**Table 1:**
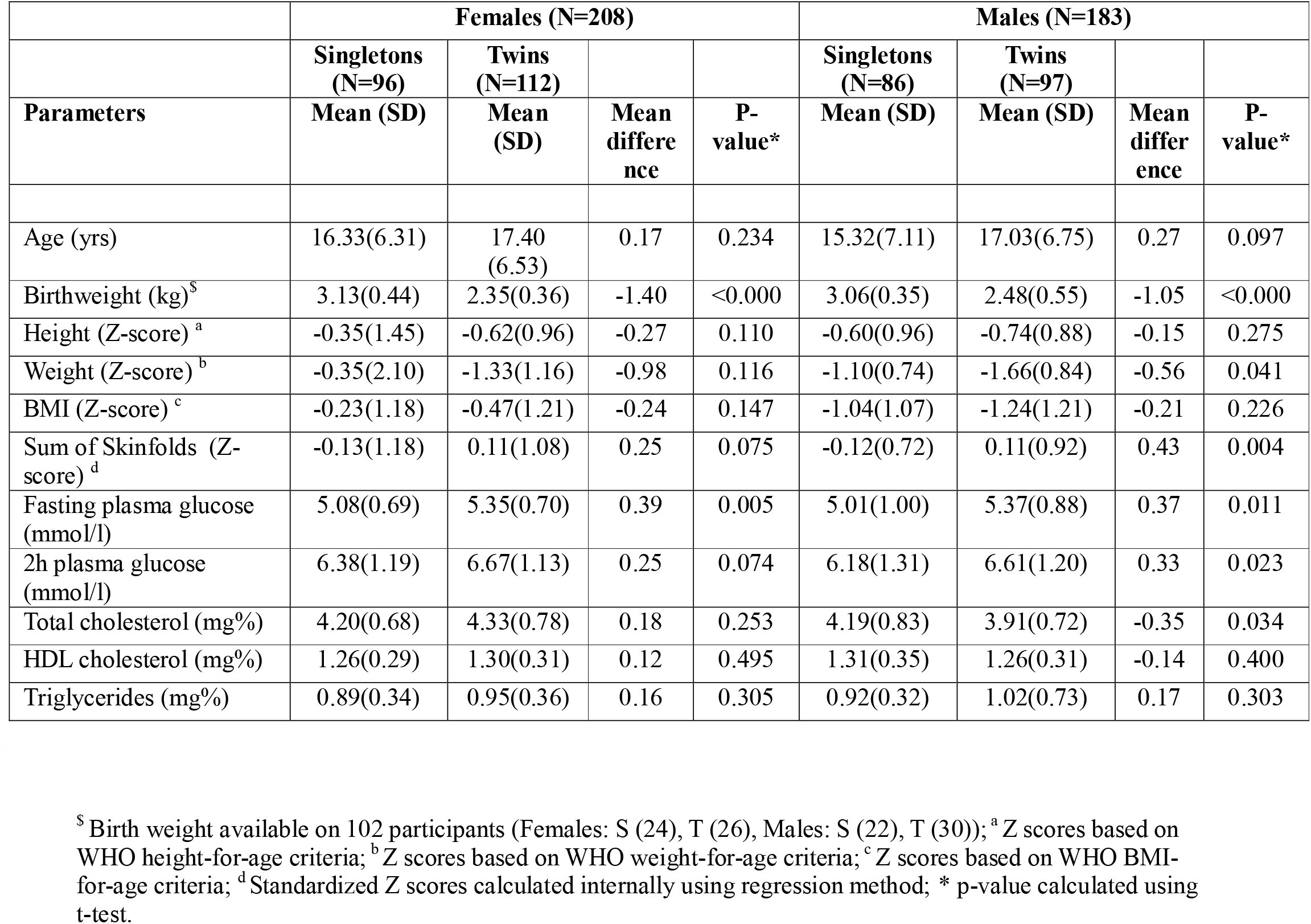
Comparison of singletons and twins.

We performed linear mixed effect model using the mean of anthropometric measurement as outcome. Exposures tested in the model included twin status (twin vs singleton), sex (males vs females), age in years, and interaction between age and sex as fixed effects. Further, two indicator variables indicating whether age is more than 8y and 16y respectively and their interaction was also included as fixed effects, to adjust for the additional effect of age at these growth points. These age indicators were selected based on biological growth transitions: 8y for pre-pubertal and 16y for late pubertal. Random intercepts for monozygotic (MZ) and dizygotic (DZ) twins were included to account for correlations between the twins.

Let, *S* identify twins (1 for all twins, even those twins that do not have their sibling, 0 for birth singletons) and *F* identify families. Further, assume *M* identifies MZ twin pairs and *D* identifies DZ twin pairs. Let *Y* denote the outcome (height, BMI and skinfolds). For any given outcome, we allow MZ twin pairs to have some covariance different from DZ twin pairs, which can be achieved by assuming a random interecept model for each level of *M* and *D*. Let, *m* be the random intercept associated with *M* and *d* be the random intercept associated with *D*, the model for the *i*^th^ subject is given by,

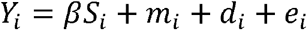

where *e* is the error term independent of *d* and *m*. We assume the random effects to follow a Gaussian distribution with 0 mean and positive variance independent of each other. The main parameter of interest is *β* (the twins effect). We used R package lme4 ^22^ to fit the random effect models. Further details of fitting the model are provided in supplementary material (Appendix S1). This model was used to predict the values given in the table.

We investigated the association of twin status (twin vs singleton) with fasting and 2-hr glucose concentrations by comparing the two groups by t-test (table 1). We further investigated if differences in age, sex, BMI and skinfolds between two groups influenced the difference (ANCOVA).

For ease of interpretation, results in tables and figures are presented using native variables. For formal statistical analysis, we used log-transformed variables. All statistical analysis was performed using R software (version 4.1) ^23^.

## RESULTS

The analysis comprised 209 twins (81 pairs and 47 individuals where only one of the twins was available) and 182 singletons (Table 1). Of the twins, 11 were MZ and 56 DZ twin pairs, and 14 pairs with no established zygosity. Birth weight was available on 102 participants (56 twins and 46 singletons) who were born in health institutions, these participants were younger (12.8 ± 4.8 y vs 18.8 ± 6.4 y, p<0.001) compared to those on whom birthweight was not available. Twins had lower birthweight compared to singletons (2.42 ± 0.4 vs 3.10 ± 0.5 kg, p<0.001).

At the time of study, both male and female twins were shorter, lighter, had lower BMI but higher skinfolds (adiposity) compared to singletons (table 1). To understand the anthropometric trajectory, we used the LME models to predict the outcomes at ages 8y, 12y, 16y and 20y (table 2). These ages broadly represent pre-pubertal, early and late pubertal, and young adulthood and are shown for depicting average cohort representative. There was the expected increase in height, BMI and skinfolds with increasing age. Females were shorter, had higher BMI and skinfolds compared to males. The figures 1*a*-*c* compare the anthropometry by age in twins and singletons and show that twins were shorter, had a lower BMI and higher sum of skinfolds across the whole age range compared to singletons. Twins also had higher skinfolds for each BMI compared to the singletons (Fig. 1*d*).

**Table 2:**
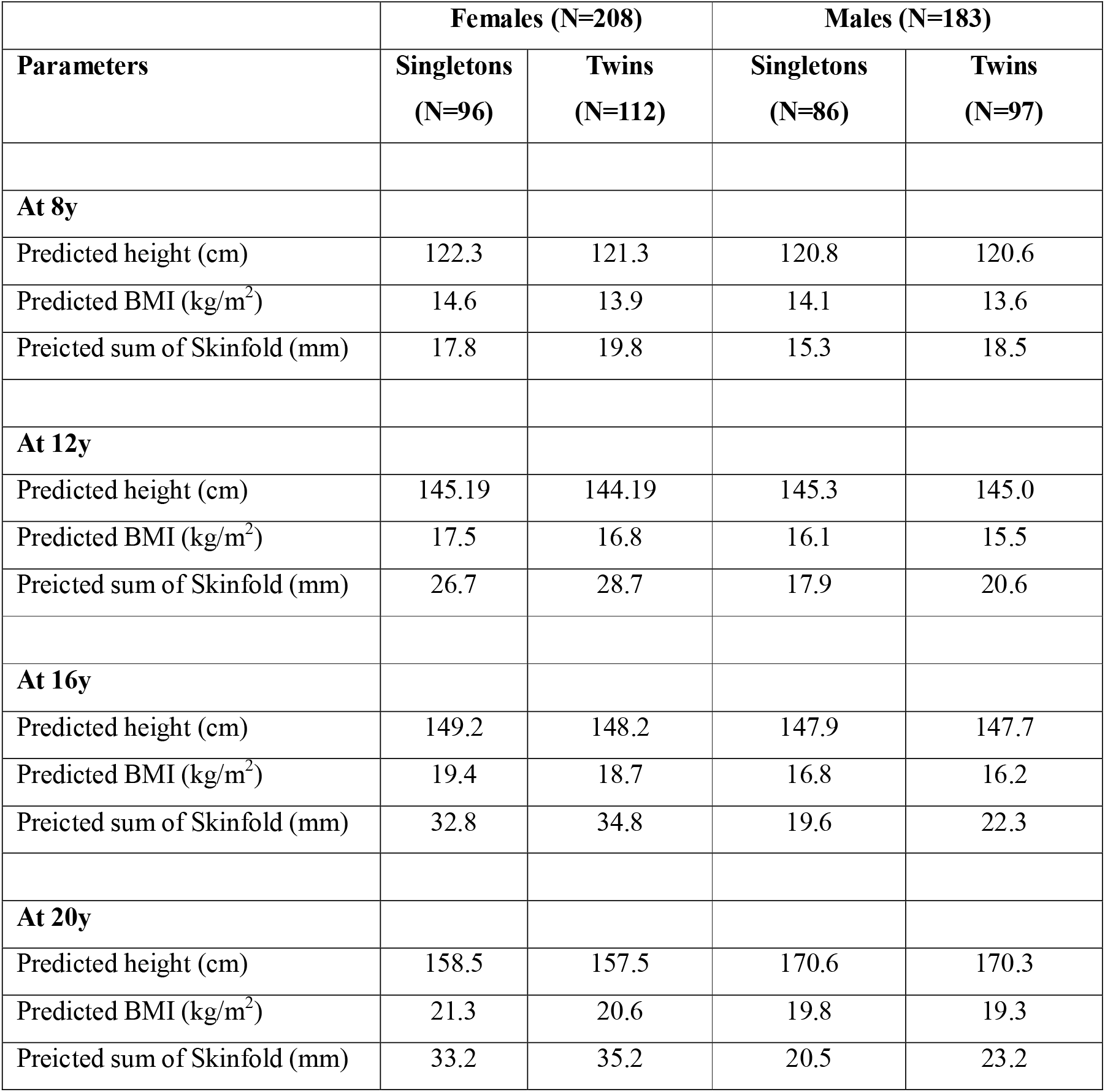
Predicted values (linear mixed effect model) between singletons and twins at different ages.

**Figure 1.**
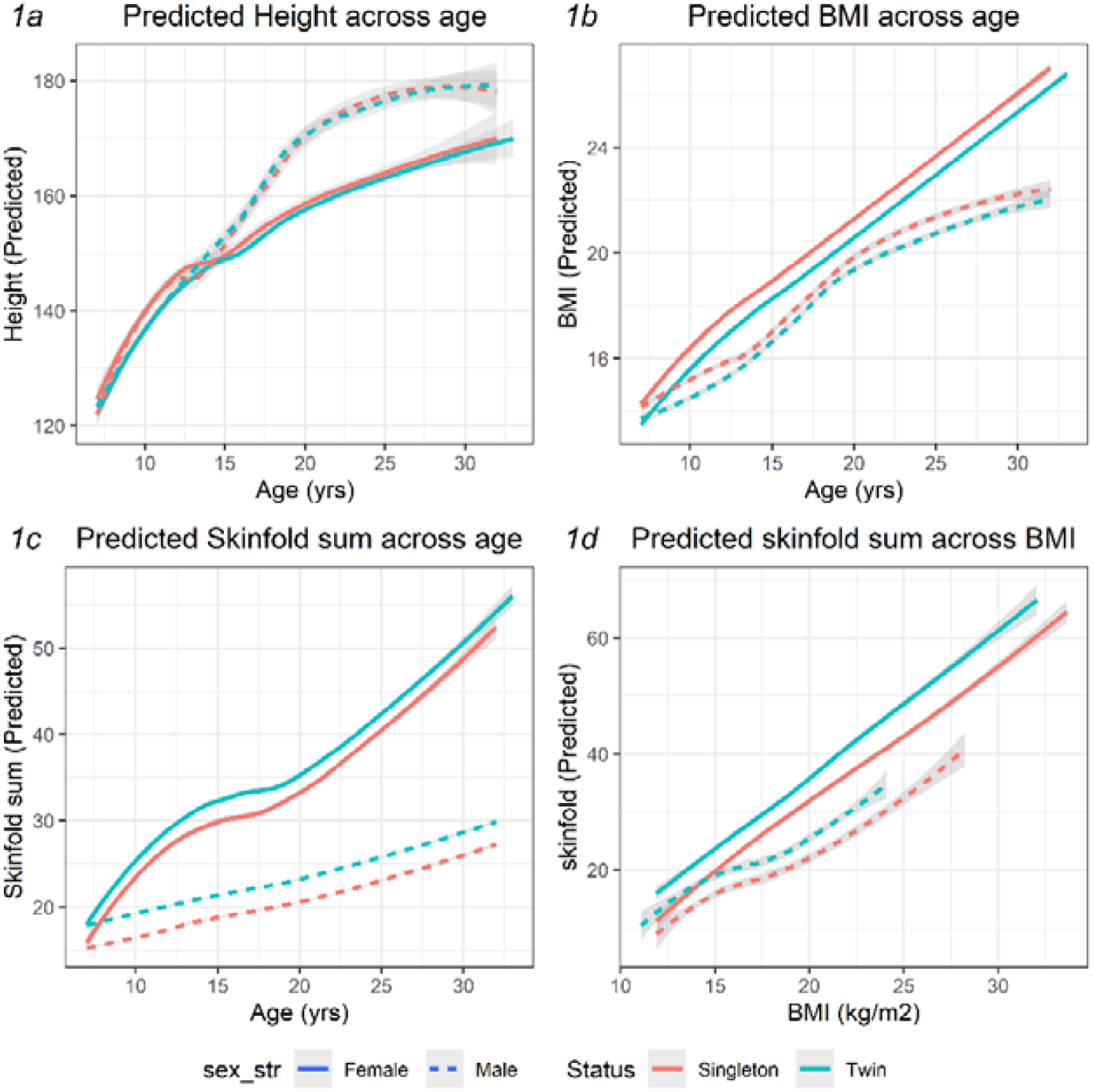
Comparison of anthropometry and body composition between singletons and twins. Solid line: Females, Dotted line: Males, Orange: Singleton, Green: Twin.

As previously reported, fasting and 2-hr plasma glucose concentrations during OGTT were higher in the twins compared to the singletons, adjusting for the age and sex difference. The difference in fasting and 2-hr plasma glucose between twins and singletons remained significant after adjusting for BMI and skinfolds. However, adjusting for birth-weight reduced the size of the difference, suggesting that birth-weight difference in the two groups might contribute to this association, though the number in this model was much lower (Supplementary table S1). There was no difference between twins and singletons in total and HDL cholesterol and triglyceride concentrations (Table 1).

## DISCUSSION

We explored the presence of a ‘thin-fat’ body composition in twins as compared to singletons using data from the Guinea-Bissau twin registry. We found that twins were thinner and more adipose than their singleton counterparts, across the age range studied. Importantly, the thin-fat twins had higher plasma glucose concentrations compared to the singletons, implying increased future diabetes risk. This was at least partially attributable to lower birthweight of twins. These findings in a novel situation (twins in a low socio-economic African country) support the role of the intra-uterine undernutrition in influencing body composition and NCD risk, first described in Indians.

‘Thinness’ refers to lower BMI and lean mass, and ‘adiposity’ to higher body fat measurements (skinfold thicknesses). This combination was considered a typically Indian (ethnic) characteristic. However, it has since been described in other South-East Asian populations from Singapore ^13^. The phenotype could have a genetic basis but no specific genetic markers have been described as yet. It is thought to be predominantly related to multigenerational maternal-fetal undernutrition of both macro- and micronutrients which would have epigenetic influence on gene expression during crucial periods of intrauterine development ^24,25^. Multigenerational undernutrition in Indians and South-East Asians is highlighted by the fact that between 1830 to 1980, these populations did not gain in height while Europeans gained ∼15 cms ^26^. The ‘thin-fat’ phenotype adds a body compositional dimension to the ‘thrifty’ phenotype described by Hales and Barker based on the association of lower birthweight with higher risk of diabetes. The relative adiposity is also reflected in biochemical (hyperglycemia) and endocrine (high leptin and insulin, and low adiponectin concentrations) abnormalities in the cord blood, further expanding the scope of the phenotype ^27^. Maternal obesity and diabetes have been shown to exaggerate the phenotype in migrant Indian populations ^28^.

The increased risk of diabetes in the ‘thin-fat’ individuals appears multifactorial. Adiposity would influence risk factors for diabetes from early age including insulin resistance ^29,30^. Lower muscle mass would also contribute to insulin resistance through its role in glucose disposal ^31^. Post-mortem studies of Indians have shown smaller organs (pancreas, liver, kidneys, etc) compared to those in Europeans, highlighting a lower ‘capacity’ in relation to higher metabolic ‘load’ ^32^. Such factors may explain failure of pancreatic beta cell insulin secretion at a young age and low BMI not only in Indians ^33,34^, but also in the twins compared to singletons in Guinea-Bissau.

The limited data on body composition of twins in the literature is mainly from the well-nourished European populations. In one study, twins had higher abdominal obesity (waist-hip ratio), insulin resistance (HOMA and Insulin Sensitivity Index) and increased prevalence of type 2 diabetes in adult life as compared to singletons ^35^. Another study which measured adiposity (DXA) in young and elderly MZ twins and a small number of singleton controls reported no significant difference between twins and singletons ^36^. The increased risk of diabetes was also not observed in a large twin registry ^37^. Thus, the strength and consistency of evidence linking lower birthweight in twins with the risk of diabetes and adiposity remains less conclusive than that in singletons from the developed countries.

Guinea-Bissau is a low-income country (Human Development Index: 0.480, Rank: 175 /189) with political instability and high prevalence of undernutrition ^38,39^. At the same time, African populations have a more muscular body composition compared to Europeans ^40,41^. Demonstration of a relatively adipose phenotype in twins compared to singletons provides support for intrauterine programming of adverse body composition by nutritional compromise. Our data showed that the twins had the expected lower birth weight (∼700 gm lower) than in singletons and were shorter, thinner (lower BMI) but fatter (higher skinfolds) at follow-up. Thus, they were ‘thin-fat’ compared to singletons and had higher glycemia. Lower birthweight explained some of the variance of higher glycemia compared to singletons, the possible reasons for which are discussed. Animal model of multigenerational maternal undernutrition has described similar findings ^42^. Sonographic measurements have usually stressed that growth in twins falters in the third trimester of gestation which is ascribed to fetal undernutrition but recent studies have demonstrated earlier growth faltering, especially in the abdominal circumference but with preservation of head circumference ^43,44^. Earlier growth failure suggests that factors in addition to under-nutrition may contribute, for example hormonal and epigenetic ^43^. Animal experiments involving fetal reduction also seem to support factors in addition to nutritional deprivation ^43^. A post-mortem study in aborted human fetuses showed that adipose tissue is demonstrable from late first trimester, their precursors divide, differentiate and mature upto 22-23 weeks of gestation, and subsequently grow only in size (hypertrophy) ^45^. It would be interesting to decide if the adiposity of twins is hyperplastic or hypertrophic or both. The developmental fate of the cells is mostly decided at gastrulation but currently there are no non-invasive tests to investigate these molecular-cellular mechanisms in human pregnancies.

Strengths of our study are a unique and rarely studied West-African low socio-economic population with high rates of twinning. Data is available on a sizeable number of twins and singletons which facilitates the comparison. Measures of skinfolds were available in addition to the usually measured weight and height. The study has limitations, being an exploratory secondary analysis. A small number of MZ twin pairs and lack of zygosity information on 14 pairs prevented us from using the classic approach of defining ‘heritability’ in twin studies. While it is tempting to attribute the phenotype to intra-uterine undernutrition, we are limited in our arguments that birthweight was available only in a fourth, and little information was available on factors such as socio-economic condition, shortened gestation and higher rates of perinatal infections and other maternal morbidities (e.g., HIV, tuberculosis, malaria) that may have contributed. In this population, twins have a 1.7-fold higher prematurity rate, compared to singletons ^18^. Relative resource deprivation would be perpetuated even after birth for twins, such as sharing of lactation as well as economic burdens on the family hampering nutrition. We were not able to directly measure or control for many of these factors in this retrospective analysis. Thus, our report may be considered only hypothesis generating, and would hopefully prompt future prospective studies which will allow careful matching to test the relative contributions of all these factors to the fetal phenotype and future disease risk.

Our results show a robust finding that African twins in Guinea Bissau are thinner but fatter compared to singletons, in a low socio-economic environment with substantial maternal undernutrition. While the data doesn’t allow us to use conventional approaches of twin analysis (separating genetic from environmental contributions by comparing MZ and DZ twins) our results should invite larger twin registries to look at this novel possibility of fetal programming of body composition. Twin studies in malnourished populations could add to the armamentarium of DOHaD researchers in the study of mechanistic aspects of fetal programming.

## Data Availability

Data is available with Dr Morten Bjerregard-Anderson for sharing to confirm our findings and for additional analyses by applying to the corresponding author with a 200-word plan of analysis. Data sharing is subject to Ethical Committee in Guinea-Bissau and the Central Ethical Committee in Denmark.

## ACKNOWLEDGEMENTS

The authors would like to thank all the participants in Guinea-Bissau, as well as the local field assistants and laboratory technicians from the Bandim Health Project, particularly Moises Soares Gomes, Leontina Indeque and Lita Indeque. The authors would also like to thank Lone Hansen, laboratory technician at the Department of Endocrinology, Odense University Hospital, Denmark, for assisting with the OGTT training and supply, as well as the biochemical analyses on collected samples. The authors would also like to thank prof. Peter Aaby, Prof. Christine Stabell Benn and Prof. Henning Beck-Nielsen for support during the project. Finally, the authors would like to acknowledge the late diabetes specialist Dr. Luis Carlos Joaquím, (who sadly died in 2012), for his assistance.

## FINANCIAL SUPPORT

The principal investigator (MBA) received a combined grant from Forsknings-og Innovations Styrelsen, the University of Southern Denmark, and Odense University Hospital. Den Danske Forsknings Fond, Odense University Hospital, Fonden for Lægevidenskabens Fremme and Aase and Ejnar Danielsen’s Foundation supported the data collection. CSY was a visiting professor to the University of Southern Denmark and Danish Diabetes Academy during this work, supported by Novo Nordisk Fonden. The funding sources had no role in the study design, data collection, data analysis or in writing of this manuscript.

## CONFLICTS OF INTEREST

The Authors declare no conflicts of interest.

## ETHICAL STANDARDS

The investigations were approved by the Ethical Committee in Guinea-Bissau. Consultative approval was obtained from the Central Ethical Committee in Denmark. Written consent (either signature or fingerprint) was obtained in all cases. For individuals <15 years consent was obtained from the mother or another caretaker.

## Notes

### Competing Interest Statement

The authors have declared no competing interest.

## References

1. Hales CN, Barker DJ. Type 2 (non-insulin-dependent) diabetes mellitus: the thrifty phenotype hypothesis. Diabetologia. 1992;35(7):595–601. doi:10.1007/BF00400248

2. Hales CN, Barker DJ, Clark PM, et al. Fetal and infant growth and impaired glucose tolerance at age 64. BMJ. 1991;303(6809):1019–1022. doi:10.1136/bmj.303.6809.1019

3. Whincup PH, Kaye SJ, Owen CG, et al. Birth weight and risk of type 2 diabetes: a systematic review. JAMA. 2008;300(24):2886–2897. doi:10.1001/jama.2008.886

4. Fall CH. Non-industrialised countries and affluence. Br Med Bull. 2001;60:33–50. doi: 10.1093/bmb/60.1.33.

5. Yajnik CS, Fall CH, Coyaji KJ, et al. Neonatal anthropometry: the thin-fat Indian baby. The Pune Maternal Nutrition Study. Int J Obes Relat Metab Disord. 2003;27(2):173–180. doi:10.1038/sj.ijo.802219

6. Yajnik CS, Lubree HG, Rege SS, et al. Adiposity and hyperinsulinemia in Indians are present at birth. J Clin Endocrinol Metab. 2002;87(12):5575–5580. doi:10.1210/jc.2002-020434

7. Modi N, Thomas EL, Uthaya SN, Umranikar S, Bell JD, Yajnik C. Whole body magnetic resonance imaging of healthy newborn infants demonstrates increased central adiposity in Asian Indians. Pediatr Res. 2009;65(5):584–587. doi:10.1203/pdr.0b013e31819d98be

8. Yajnik C. The Story of the hungry Indian foetus. NFI Bulletin. 2019; 40(3):1–8.

9. Yajnik CS, Yudkin JS. The Y-Y paradox. Lancet. 2004;363(9403):163. doi:10.1016/S0140-6736(03)15269-5

10. Lakshmi S, Metcalf B, Joglekar C, Yajnik CS, Fall CH, Wilkin TJ. Differences in body composition and metabolic status between white U.K. and Asian Indian children (EarlyBird 24 and the Pune Maternal Nutrition Study). Pediatr Obes. 2012;7(5):347–354. doi:10.1111/j.2047-6310.2012.00063.x

11. D’Angelo S, Yajnik CS, Kumaran K, et al. Body size and body composition: a comparison of children in India and the UK through infancy and early childhood. J Epidemiol Community Health. 2015;69(12):1147–1153. doi:10.1136/jech-2014-204998.

12. van Steijn L, Karamali NS, Kanhai HH, et al. Neonatal anthropometry: thin-fat phenotype in fourth to fifth generation South Asian neonates in Surinam. Int J Obes (Lond). 2009;33(11):1326–1329. doi:10.1038/ijo.2009.154

13. Deurenberg-Yap M, Schmidt G, van Staveren WA, Deurenberg P. The paradox of low body mass index and high body fat percentage among Chinese, Malays and Indians in Singapore. Int J Obes Relat Metab Disord. 2000;24(8):1011–1017. doi:10.1038/sj.ijo.0801353

14. Poulsen P, Vaag AA, Kyvik KO, Møller Jensen D, Beck-Nielsen H. Low birth weight is associated with NIDDM in discordant monozygotic and dizygotic twin pairs. Diabetologia. 1997;40(4):439–446. doi:10.1007/s001250050698

15. Phillips DI, Davies MJ, Robinson JS. Fetal growth and the fetal origins hypothesis in twins--problems and perspectives. Twin Research: the Official Journal of the International Society for Twin Studies. 2001 Oct;4(5):327-331. DOI: 10.1375/1369052012669. PMID: 11869484.

16. Hall JG. Twinning. Lancet. 2003 Aug 30;362(9385):735–43. doi: 10.1016/S0140-6736(03)14237-7. PMID: 12957099.

17. Smits J, Monden C. Twinning across the Developing World. PLoS One. 2011;6(9):e25239. doi: 10.1371/journal.pone.0025239.

18. Bjerregaard-Andersen, M., Lund, N., Jepsen, F.S. et al. A prospective study of twinning and perinatal mortality in urban Guinea-Bissau. BMC Pregnancy Childbirth. 2012; 12:140. https://doi.org/10.1186/1471-2393-12-140

19. Bjerregaard-Andersen M, Gomes GM, Hennild DE, et al. The Guinea-Bissau Twin Registry Update: A Platform for Studying Twin Mortality and Metabolic Disease. Twin Res Hum Genet. 2019;22(6):554–560.

20. Hennild DE, Bjerregaard-Andersen M, Joaquim LC, et al. Prevalence of impaired glucose tolerance and other types of dysglycaemia among young twins and singletons in Guinea-Bissau. BMC endocrine disorders 2016; 16(1): 46.

21. Bjerregaard-Andersen M, Hansen L, da Silva LI, et al. Risk of metabolic syndrome and diabetes among young twins and singletons in Guinea-Bissau. Diabetes care 2013; 36(11): 3549–56.

22. Rabe-Hesketh S, Skrondal A, Gjessing HK. Biometrical modeling of twin and family data using standard mixed model software. Biometrics. 2008;64(1):280–288. doi:10.1111/j.1541-0420.2007.00803.x

23. R Core Team (2020). R: A language and environment for statistical computing. R Foundation for Statistical Computing, Vienna, Austria. URL: https://www.R-project.org/.

24. Yajnik CS. The insulin resistance epidemic in India: fetal origins, later lifestyle, or both?. Nutr Rev. 2001;59(1 Pt 1):1–9. doi:10.1111/j.1753-4887.2001.tb01898.x

25. Yajnik CS, Yajnik PC. Fetal adiposity epidemic in the modern world: a thrifty phenotype aggravated by maternal obesity and diabetes. Am J Clin Nutr. 2020;112(1):8–10. doi:10.1093/ajcn/nqaa122

26. NCD Risk Factor Collaboration (NCD-RisC). A century of trends in adult human height. eLife 2016; 5: e13410

27. Yajnik, C. S., Lubree, H. G., Rege, S. S., Naik, S. S., Deshpande, J. A., Deshpande, S. S., Joglekar, C. V., & Yudkin, J. S. (2002). Adiposity and hyperinsulinemia in Indians are present at birth. The Journal of clinical endocrinology and metabolism, 87(12), 5575– 5580. https://doi.org/10.1210/jc.2002-020434

28. Anand SS, Gupta MK, Schulze KM, et al. What accounts for ethnic differences in newborn skinfold thickness comparing South Asians and White Caucasians? Findings from the START and FAMILY Birth Cohorts. Int J Obes (Lond). 2016;40(2):239–244. doi:10.1038/ijo.2015.171

29. Joglekar CV, Fall CH, Deshpande VU, et al. Newborn size, infant and childhood growth, and body composition and cardiovascular disease risk factors at the age of 6 years: the Pune Maternal Nutrition Study. Int J Obes (Lond). 2007;31(10):1534–1544. doi:10.1038/sj.ijo.0803679

30. Bavdekar A, Yajnik CS, Fall CH, et al. Insulin resistance syndrome in 8-year-old Indian children: small at birth, big at 8 years, or both?. Diabetes. 1999;48(12):2422–2429. doi:10.2337/diabetes.48.12.2422

31. Abdul-Ghani MA, DeFronzo RA. Pathogenesis of Insulin Resistance in Skeletal Muscle. Benian GM, editor. J Biomed Biotechnol. 2010;2010:476279. Available from: https://doi.org/10.1155/2010/476279

32. Wells, J. C., Pomeroy, E., Walimbe, S. R., Popkin, B. M., & Yajnik, C. S. (2016). The Elevated Susceptibility to Diabetes in India: An Evolutionary Perspective. Frontiers in public health, 4, 145. https://doi.org/10.3389/fpubh.2016.00145

33. Staimez LR, Weber MB, Ranjani H, Ali MK, Echouffo-Tcheugui JB, Phillips LS, Mohan V, Narayan KM. Evidence of reduced β-cell function in Asian Indians with mild dysglycemia. Diabetes Care. 2013 Sep;36(9):2772–8. doi: 10.2337/dc12-2290.

34. Yajnik CS, Bandopadhyay S, Bhalerao A, et al. Poor In Utero Growth, and Reduced β-Cell Compensation and High Fasting Glucose From Childhood, Are Harbingers of Glucose Intolerance in Young Indians. Diabetes Care. 2021;44(12):2747–2757. doi:10.2337/dc20-3026

35. Poulsen P, Grunnet LG, Pilgaard K, et al. Increased risk of type 2 diabetes in elderly twins. Diabetes. 2009;58(6):1350–1355. doi:10.2337/db08-1714

36. Malis, C., Rasmussen, E. L., Poulsen, P., Petersen, I., Christensen, K., Beck-Nielsen, H., Astrup, A., & Vaag, A. A. (2005). Total and regional fat distribution is strongly influenced by genetic factors in young and elderly twins. Obesity research, 13(12), 2139–2145. https://doi.org/10.1038/oby.2005.265

37. Petersen I, Nielsen MM, Beck-Nielsen H, Christensen K. No evidence of a higher 10 year period prevalence of diabetes among 77,885 twins compared with 215,264 singletons from the Danish birth cohorts 1910-1989. Diabetologia. 2011;54(8):2016–2024. doi:10.1007/s00125-011-2128-2

38. United Nations Development Programme 2019; Human Development Indicators.

39. World Food Programme. Guinea-Bissau Country Brief; July and August 2021.

40. Wagner DR, Heyward VH. Measures of body composition in blacks and whites: a comparative review. Am J Clin Nutr. 2000;71(6):1392–1402. doi:10.1093/ajcn/71.6.1392

41. Silva AM, Shen W, Heo M, et al. Ethnicity-related skeletal muscle differences across the lifespan. Am J Hum Biol. 2010;22(1):76–82. doi:10.1002/ajhb.20956

42. Hardikar AA, Satoor SN, Karandikar MS, et al. Multigenerational Undernutrition Increases Susceptibility to Obesity and Diabetes that Is Not Reversed after Dietary Recuperation. Cell Metab. 2015;22(2):312–319. doi:10.1016/j.cmet.2015.06.008

43. Muhlhausler, B. S., Hancock, S. N., Bloomfield, F. H., & Harding, R. (2011). Are twins growth restricted?. Pediatric research, 70(2), 117–122. https://doi.org/10.1203/PDR.0b013e31821f6cfd

44. Hiersch L, Okby R, Freeman H, et al. Differences in fetal growth patterns between twins and singletons. J Matern Fetal Neonatal Med. 2020;33(15):2546–2555. doi:10.1080/14767058.2018.1555705

45. Poissonnet, C. M., Burdi, A. R., & Bookstein, F. L. (1983). Growth and development of human adipose tissue during early gestation. Early human development, 8(1), 1–11. https://doi.org/10.1016/0378-3782(83)90028-2

