## Supplementary Material for "Twins in Guinea-Bissau have a ‘thin-fat’ body composition compared to singletons"

**Supplementary table S1: Difference between the twins compared to singletons in relation to Plasma glucose concentrations.**

|  | **F-statistics** | **P-value** |
| --- | --- | --- |
| **Fasting plasma glucose** |  |  |
| **Model 1 (N=389)** |  |  |
| Status (singleton/ twin) | 15.29 | **<0.001** |
| Age | 0.164 | 0.685 |
| Sex | 0.000 | 0.983 |
| **Model 2 (N=387)** |  |  |
| Status (singleton/ twin) | 15.92 | **<0.001** |
| Age | 0.228 | 0.633 |
| Sex | 0.011 | 0.915 |
| BMI | 0.192 | 0.661 |
| **Model 3 (N=385)** |  |  |
| Status (singleton/ twin) | 15.77 | **<0.001** |
| Age | 0.229 | 0.633 |
| Sex | 0.037 | 0.848 |
| BMI | 0.084 | 0.772 |
| Skinfolds | 0.339 | 0.561 |
| **Model 4 (N=100)** |  |  |
| Status (singleton/ twin) | 5.36 | **0.023** |
| Age | 3.475 | 0.065 |
| Sex | 5.281 | **0.024** |
| BMI | 0.731 | 0.395 |
| Skinfolds | 0.236 | 0.628 |
| Birthweight | 0.000 | 0.990 |
| **2-hr plasma glucose** |  |  |
| **Model 1 (N=389)** |  |  |
| Status (singleton/ twin) | 6.33 | **0.012** |
| Age | 0.069 | 0.793 |
| Sex | 0.000 | 0.989 |
| **Model 2 (N=387)** |  |  |
| Status (singleton/ twin) | 6.26 | **0.013** |
| Age | 0.063 | 0.8026 |
| Sex | 0.009 | 0.925 |
| BMI | 0.005 | 0.944 |
| **Model 3 (N=385)** |  |  |
| Status (singleton/ twin) | 6.18 | **0.013** |
| Age | 0.067 | 0.795 |
| Sex | 0.015 | 0.902 |
| BMI | 0.017 | 0.896 |
| Skinfolds | 1.235 | 0.267 |
| **Model 4 (N=100)** |  |  |
| Status (singleton/ twin) | 2.88 | 0.093 |
| Age | 0.002 | 0.961 |
| Sex | 1.785 | 0.184 |
| BMI | 0.005 | 0.941 |
| Skinfolds | 0.873 | 0.352 |
| Birthweight | 0.007 | 0.935 |

Difference is calculated using ANCOVA; Model 1: Adjusted for age and sex; Model 2: Adjusted for age, sex and BMI; Model 3: Adjusted for age, sex, BMI and skinfolds; Model 4: Adjusted for age, sex, BMI, skinfolds and birthweight. Birthweight was available on a subset of 102.

**Supplementary table 2: Difference between those with birthweight measurement compared to those without.**

|  | **Twins** | | | **Singletons** | | |
| --- | --- | --- | --- | --- | --- | --- |
|  | **BW available**  **(N=56)** | **BW missing**  **(N=153)** | **p-value*** | **BW available**  **(N=46)** | **BW missing**  **(N=135)** | **p-value*** |
| **Age (yrs)** | 12.8 (4.8) | 18.8 (6.4) | <0.001 | 10.7 (3.7) | 17.6 (6.6) | <0.001 |
| **Sex (% male)** | 53.5 | 43.7 | 0.209 | 47.8 | 47 | 0.928 |
| **BMI (kg/m2)** | 16.7 (3.7) | 19.1 (3.8) | 0.016 | 15.9 (2.8) | 19.3 (4.3) | 0.055 |
| **Skinfold sum (mm)** | 24.2 (12.0) | 30.0 (12.3) | 0.749 | 20.0 (7.3) | 26.6 (12.9) | 0.852 |

*p-value calculated by t-test or chi-square as appropriate.

**Appendix 1: Detail explanation of Linear Mixed Effect Model**

**Using lmer (lme4) to model correlation between related participants**

Lme4 is R package for dealing with correlated data via mixed models. The model used is described below.

**Notation**

$S$ identifies twins (1 for all twins, even those twins that do not have their sibling in the dataset, 0 for birth singletons). This is the “status” variable in the dataset.

$F$ identifies families. That is, any paired twins will share the same value for $F$. Each singleton is assigned a unique $F$ value. Each unpaired twin is also assigned a unique $F$ value; that is, for modeling covariance, unpaired twins are treated as singletons. Note that for the fixed effect (for the above $S$), unpaired twins are still (appropriately) treated as a twin. This variable is called “famid” in the attached code.

$M$ identifies monozygotic twin pairs. Monozygotic twin pairs will share the same value for $M$. Everyone else (even each member of dizygotic twin pairs) will get assigned a unique value for $M$. This variable is called “mzygo.id” in the attached code.

$D$ identifies dizygotic twin pairs. Dizygotic twin pairs will share the same value for $D$. Everyone else (even each member of monozygotic twin pairs) will get assigned a unique value for $D$. This variable is called “dizygo.id” in the attached code.

**Model**

For the primary analysis, we have not considered the heritability or formal genetic models as the sample size is relatively small for these. Therefore, the most straightforward model would be to allow monozygotic twin pairs to have some covariance and dizygotic twin pairs to have some other totally independent covariance. We achieve this by specifying random intercepts for each level of $M$ and $D$. Call $m$ the random intercept associated with $M$ and $d$ the random intercept associated with $D$. The model is:

$$Y_{i}=\beta S_{i}+m_{i}+d_{i}+e_{i}$$

where $e$ is the error term. Note that for monozygotic twin pairs $m_{1}=m_{2}$ but $d_{1}\neq d_{2}$ (with the reverse for dizygotic twin pairs). Thus, for monozygotic twin pairs $cov\left( Y_{1},Y_{2} \right)=var(m)$ and for dizygotic twin pairs $cov\left( Y_{1},Y_{2} \right)=var(d)$.

The main parameter of interest is $\beta$ (the twins effect).

1. Rabe-Hesketh S, Skrondal A, Gjessing HK. Biometrical modeling of twin and family data using standard mixed model software. *Biometrics*. 2008;64(1):280-288. doi:10.1111/j.1541-0420.2007.00803.x
